# ctDNA predicts recurrence and survival in stage I and II HPV-associated head and neck cancer patients treated with surgery

**DOI:** 10.1101/2024.02.14.24302784

**Authors:** Saskia Naegele, Dipon Das, Shun Hirayama, Sophia Z. Shalhout, Hang Lee, Jeremy D. Richmon, Daniel L. Faden

## Abstract

Human papillomavirus-associated oropharyngeal squamous cell carcinomas (HPV+OPSCC) release circulating tumor HPV DNA (ctHPVDNA) into the blood which we, and others, have shown is an accurate real-time biomarker of disease status. In a prior prospective observational trial of 34 patients with AJCC 8 stage I-II HPV+OPSCC treated with surgery, we reported that ctHPVDNA was rapidly cleared within hours of surgery in patients who underwent complete cancer extirpation, yet remained elevated in those with macroscopic residual disease. The primary outcomes of this study were to assess 2-year OS and RFS between patients with and without molecular residual disease (MRD) following completion of treatment in this prospective cohort. MRD was defined as persistent elevation of ctHPVDNA at two consecutive time points, without clinical evidence of disease. The secondary outcomes were 2-year OS and RFS between patients with and without detectable MRD after surgery. We observed that patients with MRD after treatment completion were more likely to recur compared to patients without MRD, while there was no difference in recurrence rates between patients with MRD and without MRD on postoperative day 1. OS did not significantly differ between patients with MRD after surgery or treatment completion compared to patients without MRD; however, time to death was significantly different between the groups in both settings, suggesting that with a larger sample size OS would differ significantly between the groups or that the impact of MRD detection on survival is time dependent.

## Introduction

Surgery is the most common treatment for AJCC 8 stage I-II human papillomavirus-associated oropharyngeal squamous cell carcinoma (HPV+OPSCC)^1^. Most patients who undergo surgical treatment also receive adjuvant radiation or chemoradiation^2^. Prediction of residual disease (RD) following surgery, and thus the need for adjuvant therapy, is based on clinicopathologic risk factors which have poor individual prognostic capacity. Considering the generally high overall survival and low recurrence rate, biomarkers capable of detecting RD after treatment are needed to personalize care and prevent over-treatment, which results in increased morbidity and decreased quality of life.

HPV+OPSCCs release circulating tumor HPV DNA (ctHPVDNA) into the blood which we, and others, have shown is an accurate real-time biomarker of disease status^3^. In a prior prospective observational trial of 34 patients with AJCC 8 stage I-II HPV+OPSCC treated with surgery, we reported that ctHPVDNA was rapidly cleared within hours of surgery in patients who underwent complete cancer extirpation, yet remained elevated in those with macroscopic RD^4^. Here, we report the association of post-treatment ctHPVDNA with 2-year overall survival (OS) and recurrence free survival (RFS) in this prospective cohort.

## Methods

### Study Design and Enrollment

34 previously untreated patients with stage I and II HPV+OPSCC were prospectively enrolled and underwent informed consent. All patients underwent transoral robotic surgery and concurrent selective neck dissection followed by risk-adjusted adjuvant therapy determined by a multi-disciplinary tumor board. Ten milliliters of blood was collected according to the following schema: 1) pretreatment, 2) postoperative day (POD) 1, 3) POD 7, 4) POD 30, 5) every 3-6 months after treatment completion. Detailed eligibility criteria, histopathologic diagnostic criteria, adjuvant treatment schemas, and ctHPVDNA detection methodology by custom droplet digital (dd)PCR assays for HPV genotypes 16, 18, 33, 35 and 45 were previously described^4^.

### Statistical analysis

The primary outcomes of the study were to assess 2-year OS and RFS between patients with and without molecular residual disease (MRD) following completion of treatment. MRD was defined as persistent elevation of ctHPVDNA at two consecutive time points, without clinical evidence of disease. The secondary outcomes were 2-year OS and RFS between patients with and without detectable MRD after surgery. Detectable MRD after surgery was defined as >1 copy/mL on POD 1. OS was measured from the date of biopsy-proven diagnosis. RFS was measured from the date of surgery. OS and RFS rates at 2 years are reported with exact binomial 95% confidence intervals and log rank test is used to evaluate if OS and RFS are different across groups with respect to time.

## Results

33 HPV+OPSCC patients were included in the analysis (Table 1). Death occurred in 2/3 patients (66.6%) with MRD following completion of treatment and 0/29 patients (0%) without MRD. The 2-year OS was 33% (95% CI 0.8%-90.6%) for patients with MRD following treatment completion and 100% (95% CI 88.1%-100%) for patients without MRD. Log-rank test of Kaplan-Meier curves showed survival was significantly different between the groups (Figure 1A). Recurrence occurred in 3/3 patients (100%) with MRD following completion of treatment and 0/29 patients (0%) without MRD. The 2-year RFS was 0% (95% CI 0.0%-70.8%) for patients with MRD after treatment completion and 100% (95% CI 88.1%-100%) for patients without MRD. Log-rank test of Kaplan-Meier curves showed recurrence was significantly different between the groups (Figure 1B). Lead times from MRD to clinical detection of recurrence were 56, 152, and 415 days.

**Table 1.** Patient Characteristics.

| Characteristic | Longitudinal *n* (%) |
| --- | --- |
| Mean age (range) | 62 (49-85) |
| Sex |  |
| Male | 29 (87) |
| Female | 4 (12) |
| Follow up range (years) | 1.3-3.8 |
| Follow up median (years) | 2.66 |
| TNM status |  |
| T |  |
| 0 | 0 |
| 1 | 17 (52) |
| 2 | 15 (45) |
| 3 | 1 (3) |
| 4 | 0 |
| N |  |
| 0 | 4 (12) |
| 1 | 27 (82) |
| 2 | 2 (6) |
| 3 | 0 |
| M |  |
| 0 | 33 (100) |
| 1 | 0 |
| AJCC 8 Staging |  |
| 1 | 30 (91) |
| 2 | 3 (9) |
| 3 | 0 |
| 4 | 0 |
| Recurrences | 3 (9) |
| Deaths | 2 (6) |
| Post-surgical treatment |  |
| Observation | 9 (27) |
| Radiation | 15 (45) |
| CRT | 9 (27) |
| MRD post-surgery |  |
| Detected (# of patients receiving adjuvant treatment) (# of patients with recurrence) | 11 (11) (2) |
| Not detected (# of patients receiving adjuvant treatment) (# of patients with recurrence) | 22 (13) (1) |
| MRD post-treatment |  |
| Detected (# of patients receiving adjuvant treatment) (# of patients with recurrence) | 3 (2) (3) |
| Not detected (# of patients receiving adjuvant treatment) (# of patients with recurrence) | 30 (22) (0) |

**Figure 1.**
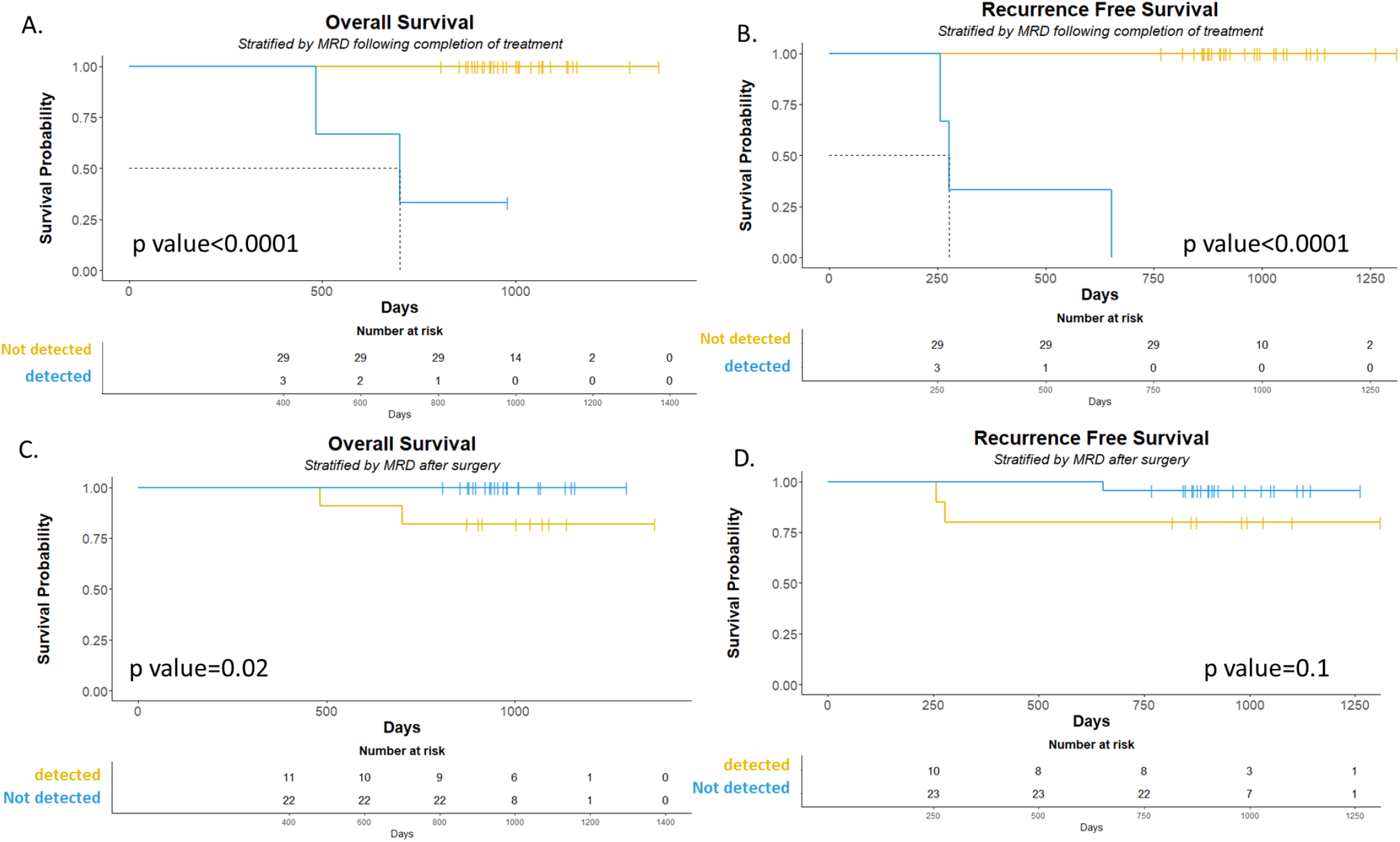
Kaplan Meier Overall Survival (A) and Recurrence Free Survival (B) curves are shown between patients with and without MRD following completion of treatment. Kaplan Meier Overall Survival (C) and Recurrence Free Survival (D) curves are shown between patients with and without MRD after surgery. (p-value, log rank test).

Death occurred in 2/11 patients (18.2%) with MRD following surgery and 0/22 patients (0%) without MRD. The 2-year OS was 81.8% (95% CI 48.2%-97.7%) for patients with MRD following surgery and 100% (95% CI 88.1%-100%) for patients without MRD. Log-rank test of Kaplan-Meier curves showed survival was significantly different between the groups (Figure 1C). Recurrence occurred in 2/11 patients (18.2%) with MRD following surgery and 1/22 patients (4.5%) without MRD. The 2-year RFS was 81.8% (95% CI 48.2%-97.7%) for patients with MRD after surgery and 95.5% (95% CI 84.6%-100%) for patients without MRD. Log-rank test of Kaplan-Meier curves showed recurrence was not significantly different between the groups (Figure 1D).

## Discussion

This is the first prospective study evaluating post-treatment ctHPVDNA as a prognostic biomarker for OS and RFS in HPV+OPSCC treated with surgery. We observed that patients with MRD after treatment completion were more likely to recur compared to patients without MRD, while there was no difference in recurrence rates between patients with MRD and without MRD on POD 1. The inferior prognostic capacity of MRD detection immediately following surgery compared to after treatment completion is likely a reflection of the high rates of adjuvant treatment, leading to ablation of residual disease. OS did not significantly differ between patients with MRD after surgery or treatment completion compared to patients without MRD; however, time to death was significantly different between the groups in both settings, suggesting that with a larger sample size OS would differ significantly between the groups or that the impact of MRD detection on survival is time dependent.

Since 66-80% of recurrences occur within the first two years of treatment, these findings highlight the importance of ctHPVDNA as a biomarker capable of predicting recurrence and survival in stage I-II HPV+OPSCC treated with surgery^5^. These findings are significant as ctHPVDNA is already being utilized clinically, both on and off trials, for decision making, yet efficacy data in the surgical setting is lacking. Limitations of this study include a large span of the CIs due to the limited size of the cohort, which in some analyses led to overlapping CIs. Despite this, large, and even complete separation events, lends high confidence that subsequent studies with greater sample sizes will reveal significant differences in all analyses with presumed large effect sizes.

## Data Availability

All data produced in the present study are available upon reasonable request to the authors.

## Acknowledgments

We would like to thank the clinical teams who helped care for the patients included in this study at Massachusetts Eye and Ear and Massachusetts General Hospital. We would also like to thank additional members of the Faden Lab who assisted with this work including Natalia Queenan, Julia Mendel, and Connor O’Bolye.

